# Collecting Mental Health Services Information from Institutions of Higher Education to Link with Student Mental Health: A Successful Pilot

**DOI:** 10.1101/2025.07.03.25330219

**Authors:** James Aluri, Brenda Vyletel, Erin Voichoski, Amelia Mercado, Jessi Haddad, Haley Henry, Julia Bell, Ashley Johnston, Aubrey DeVinney, Daniel Eisenberg, Sarah Lipson

## Abstract

**Background:** Few studies have evaluated student mental health services at institutions of higher education. Generating datasets that link institutional health services data to mental health data of students at those institutions will be a key step forward in identifying evidence-based best practices.

**Objective:** To pilot the linking of data collected by an established national student mental health survey with data collected about institutional mental health services.

**Methods:** Institutional representatives who enrolled their student body in the Healthy Minds Survey (HMS) during the 2024-2025 academic year and who did not opt out (n = 111) were invited to complete a short survey. The survey queried policies on limits for the number of mental health sessions a student could access through their campus mental health services. Institutional characteristics for HMS schools were obtained from the Integrated Postsecondary Education Data System (IPEDS).

**Results:** In total, 77 of 111 eligible institutions (69%) responded to the survey. No institutional characteristic was significantly associated with response.

**Conclusion:** This pilot demonstrated that partnering with existing research infrastructure to collect institutional mental health services data can achieve high response rates. Using existing research networks to collect institutional data and link them to student data represents a way forward for research on college mental health services.

## Introduction

Rising symptoms of mental illness among college students have been an increasing concern over the past decade.^1^ The large number of US college students (nearly 20 million)^2^ and the economic importance of higher education^3,4^ make student mental health a public health priority. Most institutions of higher education offer mental health services to their students.^5^ These clinics play an important role in addressing students’ mental health needs, but best practices for structuring, staffing, and financing of campus clinics have not been established.

Additional research^1^ is needed in this area, and this research will need to account for the heterogeneity among institutions of higher education.^6^

Generating the appropriate data to answer research questions about best practices for campus mental health services would advance the field from description to evaluation. Starting such a research endeavor faces challenges with coordinating participation, standardizing data, storing data securely, analyzing the data, and funding the time and effort needed for each of those steps. An additional challenge faced by all data collection efforts is low response rates, with one prior survey about campus mental health services achieving only a 16% response rate among institutional leaders.^7^

An alternative is to use existing research infrastructure from ongoing national studies of student health and mental health. This manuscript describes an innovative process developed to collect mental health services data from institutional leaders for linkage with the de-identified student data collected by the Healthy Minds Study (HMS). The Healthy Minds Study is a large, US survey that assesses students’ mental health at institutions of higher education that choose to participate in the survey.

## Methods

### Study Design

This cross-sectional study gathered data from institutional representatives who served as contacts for enrolling their institutions in HMS during the 2024-2025 academic year (AY). Data collection occurred between February and May 2025. The HMS is reviewed and approved by Advarra’s Institutional Review Board (IRB), and the studies methods have been described in elsewhere.^8,9^ The primary data collection from institutional leaders, linkage to the de-identified student data, and analysis of the de-identified linked dataset were reviewed and approved by the IRB of the Johns Hopkins University (JHU) School of Medicine.

### Recruitment and Sample

All institutional representatives who enrolled their institution in HMS during the 2024-2025 AY were contacted by the HMN research team, informed of the present study, and given a chance to opt out. Many representatives held leadership positions at their institution (e.g., Vice Provost, or Director of a Health Clinic). Following the approved protocol, the name, institution, and email address of leaders who did not opt out (n = 114) were then provided to the JHU research team. The JHU team emailed those contacts introducing the purpose of the research and inviting them to participate in the separate study. During the study, three leaders indicated that they were no longer planning to administer the HMS student survey and were no longer eligible for the study. HMS contacts who felt they were not well positioned to provide institutional information about session limits at their campus mental health services were given the option to refer an appropriate participant; eight HMS contacts referred the research team to one of their colleagues. Non-respondents were contacted with up to eight additional reminders to participate.

### Measures & data collection

All eligible participants were invited to participate in an online Qualtrics survey that included a maximum of nine questions about session limits. Each participant was instructed to enter their institutional information using an institutional linking ID provided by the HMN research team. The JHU research team was blinded to the institutional identities associated with each linking ID to preserve the de-identified nature of the information provided by institutions. Each participant was provided with a $20 Amazon gift card as compensation for their time.

Collection of the institutional mental health services data was concluded in May 2025. The HMS collection of student responses also ended in May 2025. The HMN research team will use the linking IDs to link data from institutions to de-identified data from students at those institutions.

Institutional characteristics were obtained from the Integrated Postsecondary Education Data System (IPEDS), with sector (public or private), size (categorized into < 1,000; 1,000-19,999; and 20,000 or more), and type (four-year school versus two-year school). Four-year schools offered bachelor’s degrees or higher, while two-year schools were limited to offering associates degrees or comparable certificates requiring at least two years. Two-year schools are often referred to as community colleges.

### Analysis

Response rates were calculated using the total number of eligible institutions in a category divided by the number who participated in that category. Chi-square tests were used to test for significant differences in the distribution of respondents by institutional characteristic.

## Results

In our sample of 111 colleges and universities (see Table 1), most institutions were public (n = 66) and four-year schools (n = 82). Of the five enrollment size categories, most institutions (n = 88) were located in the middle three (1,000-4,999; 5,000-9,999; and 10,000-19,999 students). In total, 77 of 111 eligible institutions (69%) responded to the survey evaluating session limits at campus mental health services. No institutional characteristic was significantly associated with response.

**Table 1:**
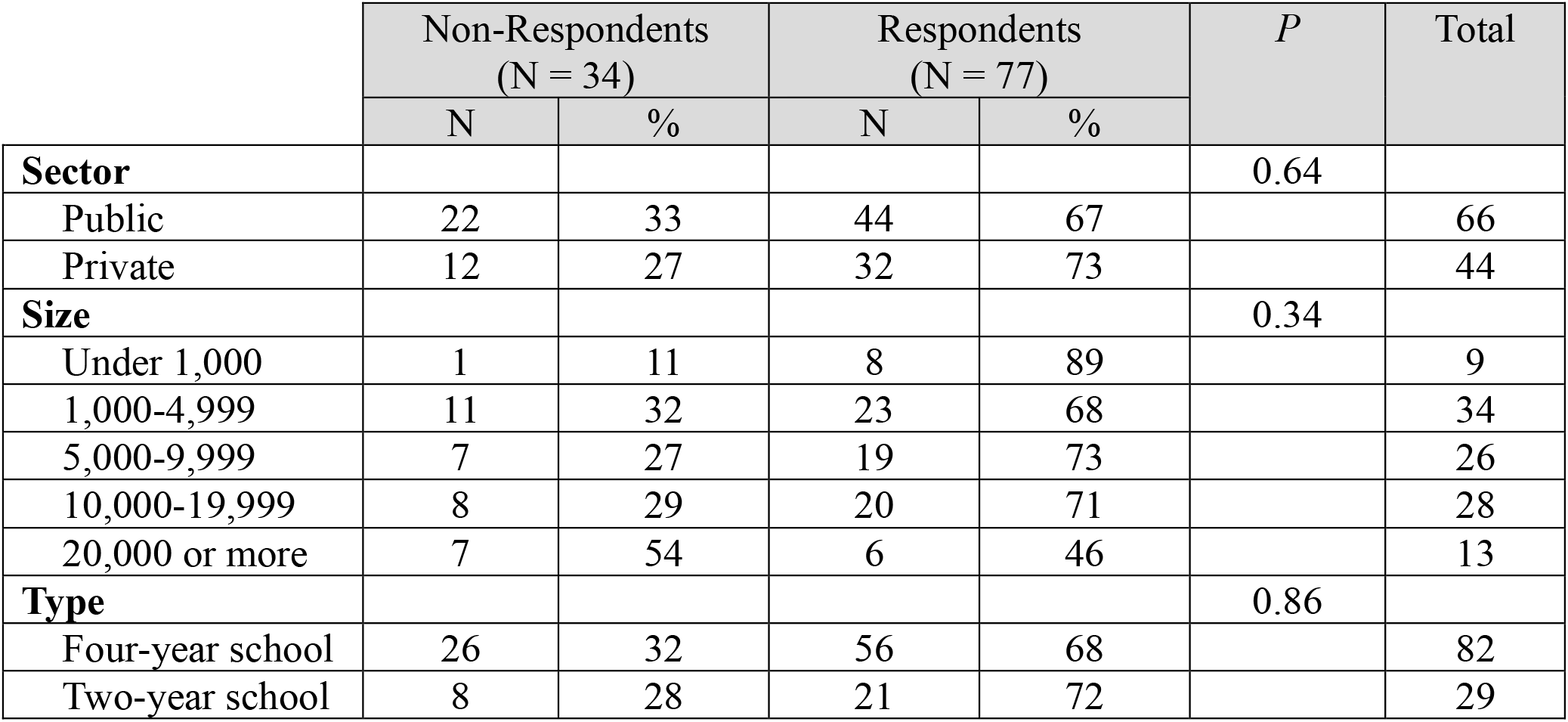
Survey Responses, Stratified by Institutional Sector, Size, and Type.

## Discussion

An innovative partnership between the existing Healthy Minds team and an external research team successfully collected focused data on campus mental health services from 77 institutions (69% of eligible institutions) while maintaining confidentiality of the student survey data.

This study’s 69% response rate surpasses the average response to online surveys (44%)^10^ and the response rate to a recent study conducted by a JH-based team on campus mental health services (16%).^7^ Several factors likely contributed to the high response rate including (a) an initial explanation about the study from the Healthy Minds research coordinator who had an existing relationship with the institutional contact, (b) the ability to partner with an established brand in college mental health (HMS), (c) offering compensation for participation, and (d) regular email reminders.

This response rate analysis did not include all institutions that administered HMS during the 2024-2025 AY, which could introduce bias. A total of 21 institutions that administered the HMS were not included in the study for various reasons including enrolling in HMS after the data linkage sample had been selected and opting out of the data linkage project. Investigating why some institutions opted out and why some who did not opt out still did not respond could yield opportunities to further improve the response rate. Another limitation is that this approach relies on cross-sectional data, which cannot make strong causal claims. Still, this approach has the advantage of low costs and large samples and could be used to identify relationships that merit further scrutiny with costly, longitudinal studies.

This study examined the relationship between the limitation of sessions students can use at campus mental health clinics (session caps) and patterns in student utilization of campus mental health services. That analysis will be executed after data linkage occurs in Autumn 2025. For next steps, this process could be applied to other questions in college mental health that involve multi-institutional comparisons of differences in health services approaches.

## Conclusion

The ability to link institutional data on campus mental health services and student data on mental health service utilization characteristics represents an important advance in the field. This pilot demonstrated that partnering with existing research infrastructure to collect institutional mental health services data can achieve higher than average response rates.

## Data Availability

HMS data are available online through request at https://healthymindsnetwork.org/hms/?__cf_chl_tk=wxK99v4s3YzdK3ep1arjhSIftQW8AJPacoIlrv3tsV4-1750796006-1.0.1.1-hfZzZ7hR8PDc3UlG8WC6RpdbZYP1BQ_fWhNsKf71iN8
Institutional services data produced in the present work are stored by the authors

## Data Availability

Data from the Healthy Minds Student Survey are available from the Healthy Minds Network by request.

